# Patterns of morbidity and mortality among neonates admitted in Mirwais regional hospital, Kandahar, Afghanistan

**DOI:** 10.1101/2024.05.13.24307260

**Authors:** Zarghoon Tareen, Ahmad Tameem Tareen, Mohammad Essa Amirzada, Nik Mohammad Zafare

## Abstract

**Background:** Neonatal health is a critical global concern that reflects the national and global progress and challenges of health systems. Neonatal morbidity and mortality are major global public health problems.

**Methods:** We conducted a hospital-based cross-sectional descriptive study of all neonates admitted to the NICU of the pediatric ward of Mirwais Regional Hospital between March 2023 and February 2024.

The objective of this study was to determine the pattern of neonatal morbidity and mortality as well as to compute case fatality rates in Mirwais Regional Hospital in Kandahar City, Afghanistan.

**Results:** In our study, there were 3387 newborns admitted to the neonatal ward of Mirwais Regional Hospital over a one-year period (from March 2023 to February 2024). Prematurity was the most common reason for admission to the neonatal ward (1048/3387, 30.9 %). Other common conditions included neonatal sepsis (909/3387, 26.8%), birth asphyxia (785/3387, 23.1%), jaundice (466/3387, 13.7%), and tetanus (9/3387, 0.3%). Surgical causes contributed to (170/3387, 5.0%) morbidities.

The overall mortality rate was 13.0% (442/ 3387). The major causes of mortality were premature birth (39.8%, 176/442), birth asphyxia (25.6%, 113/442), and neonatal sepsis (22.1%, 98/442). Other causes of mortality were neonatal jaundice (7.9%, 35/442), neonatal tetanus (0.9%, 4/442), and surgical disease (3.6%, 16/442). Neonatal tetanus had the highest mortality rate (44.4%).

**Conclusion:** Prematurity was the most common reason for admission, followed by birth asphyxia, neonatal sepsis, and jaundice. The overall mortality rate was 13%. Prematurity is a leading cause of neonatal mortality. Neonatal tetanus has a high mortality rate.

**Author summary:** Neonates were defined as those the in first 28 days of life. In humans, they are more vulnerable to diseases than any other age group. Neonatal morbidity and mortality are major public health problems with approximately 3.1 million babies worldwide dying each year in the first month of life. The main goal of this study was to estimate the types of neonatal morbidity and mortality in Merwais Regional Hospital, located in Kandahar City, Afghanistan. This study found that premature birth (birth before 37 weeks of gestation) was the most common reason for admission to the neonatal ward. Other common conditions include neonatal sepsis, birth asphyxia, neonatal jaundice, and neonatal tetanus. The overall mortality rate was high in the present study. The main cause of mortality was prematurity, followed by birth asphyxia and neonatal sepsis. Interventions should be planned and implemented at different levels in the community to prevent and reduce the incidence of neonatal diseases and deaths.

## INTRODUCTION

Neonatal health is a critical global concern, reflecting the national and global progress and challenges of health systems (1). Neonatal morbidity and mortality are major global public health challenges with approximately 3.1 million babies worldwide dying each year in the first month of life (2). In 2022, nearly half (47%) of all deaths in children under five years of age occurred in the newborn period (the first 28 days of life), which is among the most vulnerable periods of life and requires intensified quality intrapartum and newborn care (3). Most neonatal deaths occur in the first week of life and are preventable with equitable access to adequate, evidence-based maternal and newborn healthcare (4). Africa had the highest neonatal mortality rate in 2018, at 28 deaths per 1,000 live births, followed by Central and Southern Asia, with 25 deaths per 1,000 live births (3). Similar to other developing countries, approximately three-fourths of under-five deaths occur in the early neonatal period in Afghanistan; this proportion is about one-third of the total under-five deaths, and it is estimated that NMR was 22 per 1000 live births in Afghanistan in 2015 (5).

While birth asphyxia, prematurity, and infections are the leading causes of adverse neonatal outcomes, congenital anomalies contribute significantly to neonatal morbidity and mortality an estimated 7.9 million children are born with major congenital anomalies every year, and the proportion of global neonatal mortality due to these defects increased from 3% in 2008 to 4.4% in 2013 (6).

A study conducted in Karachi, Pakistan, reported that the highest number of study participants had preterm birth, low birth weight, sepsis, respiratory distress syndrome, birth asphyxia, meconium aspiration syndrome, neonatal jaundice, pneumonia, hyaline membrane disease, and congenital malformations. Of the 1069 patients, 148 died (13.8%) and one-third died due to sepsis (7).

Another retrospective study conducted on neonatal case management and outcomes in rural Rwanda reported that premature birth, neonatal infection, and asphyxia were the three most common primary diagnoses. The overall mortality rate was 13.3%, and neonates with prematurity and LBW had higher mortality rates than the others (8).

A study conducted in Kenya found that the clinical diagnoses of neonatal sepsis, prematurity, neonatal jaundice, neonatal encephalopathy, tetanus, and neonatal meningitis accounted for over 75% of inpatient neonatal admissions. The study also reported that, Tetanus 256/390 (67%), prematurity 554/1,280 (43%) and neonatal encephalopathy 253/778(33%) had the highest case fatality (9)

A study was conducted in India has reported that, Hyperbilirubinemia (7%) was the leading cause of neonatal morbidity, followed by sepsis (3.99%) and respiratory distress (3.9%). Congenital malformations were observed in 1.75% of cases.

In this study, birth asphyxia (29.85%) and respiratory distress (22.38%) were the leading causes of death, followed by extreme prematurity (22.3%), and sepsis (14.9%) (7).

A study conducted in a tertiary hospital in Nigeria reported that the common morbidities were neonatal sepsis (39.3%), prematurity (16.9%), birth asphyxia (18.2), and neonatal jaundice (12.0%). Neonatal tetanus and congenital anomalies were responsible for 4.0% and 2.3% of the patients, respectively.

In this study, the highest mortality was observed in premature birth (26.1), neonatal sepsis (24.6%), and severe birth asphyxia (23.2%). Neonatal tetanus and neonatal jaundice were responsible for (13.0%) and (7.2%) respectively (10).

## MATERIALS AND METHODS

### Study design and study area

We conducted a hospital-based cross-sectional descriptive study of all neonates admitted to the NICU of the Pediatrics Ward of Mirwais Regional Hospital between March 2023 and February 2024 (Hamal 1, 1402 and Hoot29, 1402). Mirwais Regional Hospital was constructed by China in 1979 to provide health services for people from five provinces in the southwest region of Afghanistan, which is in Kandahar City, Afghanistan. The inclusion criteria were all neonates admitted to the NICU of the pediatric ward from birth to 28 days of age.

The objective of the study was to determine the pattern of neonatal morbidity and mortality as well as compute case fatality rates in Mirwais regional hospital in Kandahar city, Afghanistan.

### Ethical considerations

This study was approved by the Kandahar University Ethics Committee (code number KDRU-EC-2023.15). Only patient initials and medical registration numbers were used for data collection. Prior to entering the computer for the analysis, the collected data were coded and de-identified. Informed consent was not obtained from any patient (not applicable).

### Samples Collection

Medical records of all neonates admitted to the NICU of the pediatric ward from March 2023 to February 2024 (Hamal 1, 1402 to Hoot29, 1402) were retrieved and analyzed. A pro forma record form was designed and used to extract information from the medical records.

### Statistical Analysis

The data were analyzed using IBM SPSS version 26. In this study, we used descriptive statistics such as frequency and percentage. Instead of the names of participants by using codes and numbers, the confidentiality of the study was ensured.

## Results

### Morbidity

In our study, there were 3387 newborns admitted to the neonatal ward of Mirwais Regional Hospital over a one-year period (from March 2023 to February 2024). Prematurity was the most common reason (1048/3387, 30.9%) for admission in the neonatal ward. Other common conditions included neonatal sepsis (909/3387, 26.8%), birth asphyxia (785/3387, 23.1%), neonatal jaundice (466/3387, 13.7%), and neonatal tetanus (9/3387, 0.3%). Surgical causes contributed to (170/3387, 5.0%) morbidities, as shown in Table 1.

**Table 1:**
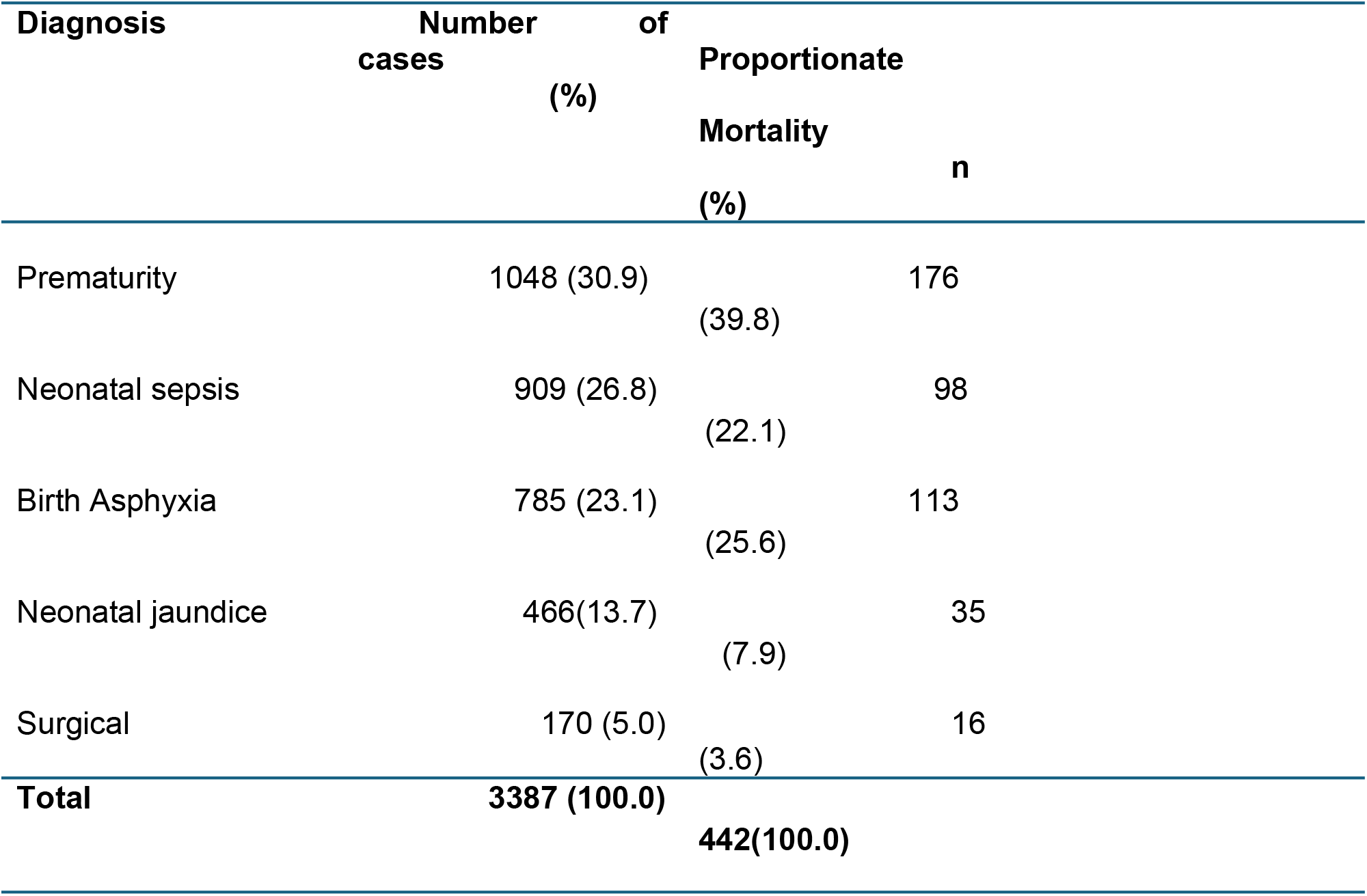
Disease pattern and mortalities.

### Mortality

The overall mortality rate was 13.0% (442/ 3387). Major causes of mortality were prematurity (39.8%, 176/442), birth asphyxia (25.6%, 113/442) and neonatal sepsis (22.1%, 98/442). Other causes of mortality were neonatal jaundice (7.9%, 35/442), neonatal tetanus (0.9%, 4/442), and surgical diseases (3.6%, 16/442) (Table 1).

### Case fatality rates

Figure 1 shows the case fatality rates in our study. Neonatal tetanus had the highest case fatality rate (44.4%), followed by premature birth (16.8%), birth asphyxia (14.4%), and neonatal sepsis (10.8%). The fatalities for neonatal jaundice and surgical diseases were 7.5% and 9.4%, respectively.

**Figure 1:**
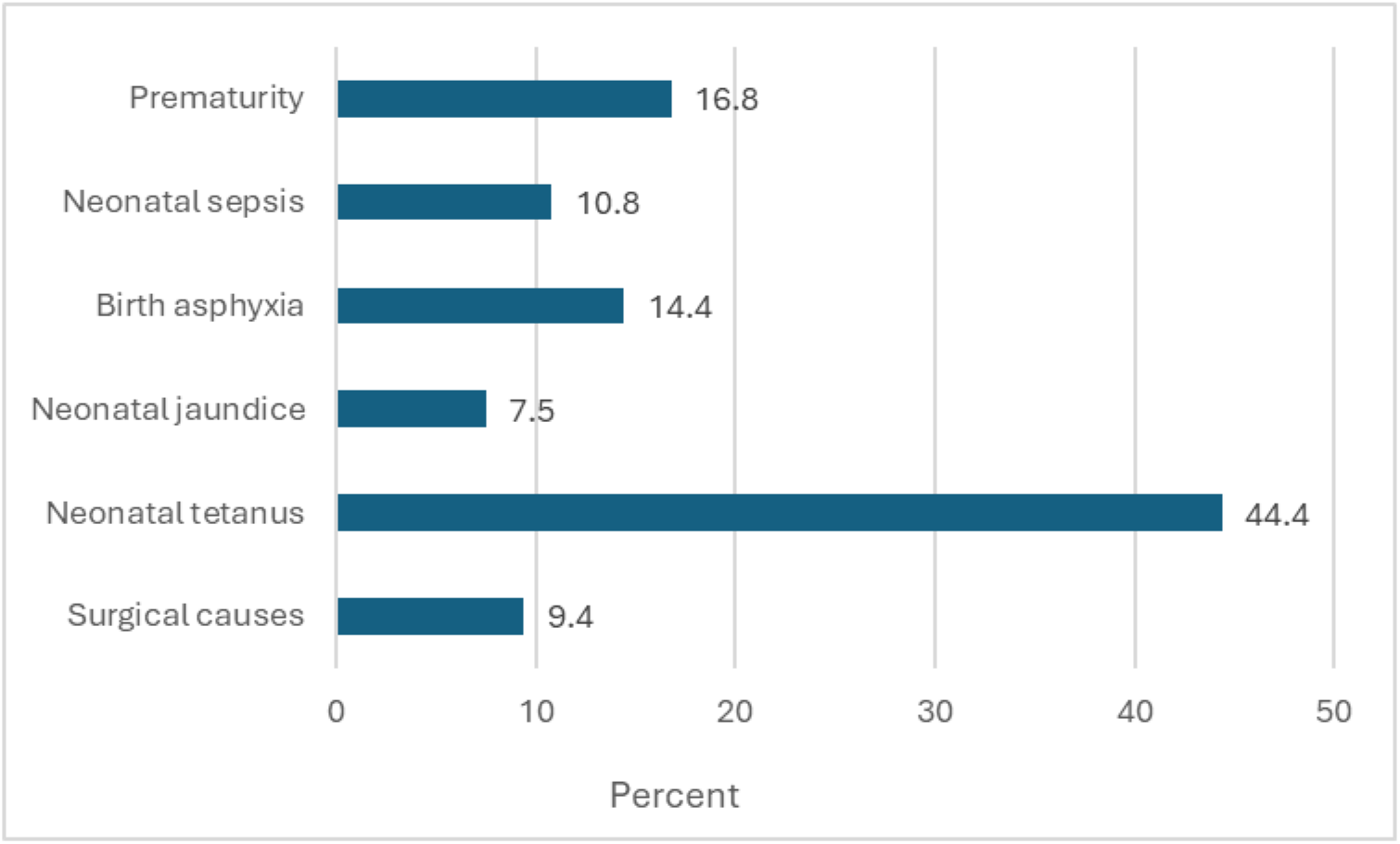
case fatality rates.

## Discussion

This study revealed the morbidity and mortality of neonatal diseases and computed the case fatality of neonatal diseases admitted to Mirwais Regional Hospital in Kandahar City, Afghanistan. In this study, the common causes of neonatal morbidities were prematurity (30.9%), neonatal sepsis (26.8%), birth asphyxia (23.1%), neonatal jaundice (13.7%), neonatal tetanus (0.3%), and surgery (5.0%). Similar patterns have been reported in several previous studies. Shahani et al. reported that the main causes of neonatal admission in Pakistan wards were sepsis (17.0%), prematurity (14.01%), birth asphyxia (10%), and neonatal jaundice (5.25%). The patterns of neonatal admissions were similar to those observed in our study (19). Another study from Pakistan reported that the main patterns of neonatal admission were prematurity (32.30%), sepsis (28.91%), birth asphyxia (11%), and neonatal jaundice (5%) (16) A study conducted in Nigeria by also et al. reported that the common causes of neonatal admission in the neonatal ward were neonatal sepsis (39.3%), prematurity (16.9%), birth asphyxia (18.2%), neonatal jaundice (12.2%), and neonatal tetanus (4%) (10). The reported patterns seems same as in our study, but the frequency of prematurity in our study was higher (30.9% vs 16.9%) and lower frequency of neonatal tetanus in this study (0.3% vs 4.0%) (10), the same result was also reported from another study from Nigeria (11)

A study conducted in India by Saini et al. showed similar results. They reported that the most common causes of neonatal morbidity were neonatal jaundice (7.0%), prematurity (4.3%), and birth asphyxia (4.0%). This result is similar to that of the present study (7). A study from South Africa reported more admissions due to sepsis, jaundice, and asphyxia (12). The patterns are different in developed countries. A study from Canada revealed that the main causes of neonatal admission were extreme prematurity, asphyxia, and congenital anomalies (13).

The overall mortality rate was 13.0% (per 100 morbidities). Several studies from different parts of the world have reported similar results. A study done by Kumar et al. reported a mortality rate of 13.6% (14). A study by Saini et al. in India reported a mortality rate of 11.3% (7). A study conducted in Nigeria by U et al. reported a similar result (13.2%) (10). A study by Ekwochi et al. reported a mortality rate of 14.2 %, which is similar to that in our study (15). A study conducted in Pakistan reported a mortality rate of 17.15% (16). A study from Bangladesh reported a slightly higher mortality rate than our study (20.6% vs. 13.0%) (17). This variation may be due to other factors and facilities available in neonatal intensive care units because neonatal survival is dependent on these factors.

In our study, prematurity was found to be a leading cause of neonatal mortality (39.8%), followed by birth asphyxia (25.6%), and neonatal sepsis (22.1%). Neonatal jaundice and tetanus were responsible for 7.9% and 0.9% of patients, respectively. Surgical causes accounted for 5% of neonatal mortality. A study by Saini et al. reported that the leading causes of neonatal mortality were birth asphyxia (29.85%), prematurity (22.38), and neonatal sepsis (15%) (7). Another study from India reported that premature birth (25.68%) was a common cause of neonatal mortality. (18). Another study done by Also U reported that premature birth, neonatal sepsis, severe birth asphyxia, and neonatal tetanus are major causes of neonatal mortality (10). A study conducted by Hussain in Pakistan reported that prematurity (26.78%) was the leading cause of neonatal mortality, followed by birth asphyxia (21.81%) and neonatal sepsis (15.22%) (16). Neonatal tetanus was associated with a high mortality rate (44.4%). This result was the same as that of a study by U (42.8%) (10). Again, these variations are due to the availability of modern facilities and trained health professionals in the neonatal wards.

## Conclusion

In this study, premature birth was the most common reason for admission, followed by birth asphyxia, neonatal sepsis, and neonatal jaundice. Overall mortality rate was 13%. In our study, we found prematurity to be the leading cause of neonatal mortality, followed by birth asphyxia and neonatal sepsis. Neonatal tetanus had high fatality rate.

## Limitations

A major limitation of this study is incomplete data, which is common in retrospective data collection. We were unable to retrieve all demographic and clinical information regarding the neonates in this study.. Furthermore, this was a hospital-based cross-sectional descriptive study, and may not be generalizable to the general population.

## Data Availability

Data will be available upon request

## Acknowledgments

We present our highest and sincere thanks to the authorities of Faculty of Medicine, Kandahar University and Kandahar Directorate of Public Health. We are also very thankful to the head, trainer specialist, and staff of the pediatric ward of Mirwais Regional Hospital.

## Author Contributions

Conceptualization: Zarghoon Tareen, Ahmad Tameem Tareen, Mohammad Essa Amirzada, Nik Mohammad Zafare.

Data curation: Zarghoon Tareeen.

Formal analysis: Zarghoon Tareen

Methodology: : Zarghoon Tareen, Ahmad Tameem Tareen

Project administration: Zarghoon Tareen, Ahmad Tameem Tareen, Mohammad Essa Amirzada

Resources: Nik Mohammad Zafare.

Software: Zarghoon Tareen.

Supervision: Zarghoon Tareen.

Validation: Zarghoon Tareen.

Visualization: Zarghoon Tareen.

Writing – original draft: Zarghoon Tareen.

Writing – review & editing: Zarghoon Tareen, Ahmad Tameem Tareen, Mohammad Essa Amirzada, Nik Mohammad Zafare.

Financial support and sponsorship

Self-sponsored study, no grants from anybody or organization.

## References

1. Ou Z, Yu D, Liang Y, He H, He W, Li Y, Zhang M, Gao Y, Wu F, Chen Q. Global trends in incidence and death of neonatal disorders and its specific causes in 204 countries/territories during 1990– 2019. BMC public health. 2022 Feb 19;22(1):360.

2. Tran HT, Doyle LW, Lee KJ, Graham SM. A systematic review of the burden of neonatal mortality and morbidity in the ASEAN Region. WHO South-East Asia Journal of Public Health. 2012 Jul 1;1(3):239–48.

3. Naji FA. Risk Factors for Malnutrition Among Children Aged 6-59 Months Attending Sos Hospital in Mogadishu, Somalia (Doctoral dissertation, University of Nairobi).

4. Nabwera HM, Wang D, Tongo OO, Andang’o PE, Abdulkadir I, Ezeaka CV, Ezenwa BN, Fajolu IB, Imam ZO, Mwangome MK, Umoru DD. Burden of disease and risk factors for mortality among hospitalized newborns in Nigeria and Kenya. PloS one. 2021 Jan 14;16(1):e0244109.

5. Kibria GM, Burrowes V, Choudhury A, Sharmeen A, Ghosh S, Mahmud A, Kc A. Determinants of early neonatal mortality in Afghanistan: an analysis of the Demographic and Health Survey 2015. Globalization and health. 2018 Dec;14:1–2.

6. Ajao AE, Adeoye IA. Prevalence, risk factors, and outcomes of congenital anomalies among neonatal admissions in OGBOMOSO, Nigeria. BMC pediatrics. 2019 Dec;19:1–0.

7. Saini N, Chhabra S, Chhabra S, Garg L, Garg N. Patterns of neonatal morbidity and mortality: A prospective study in a District Hospital in Urban India. Journal of Clinical Neonatology. 2016 Jul 1;5(3):183–8.

8. Nyishime M, Borg R, Ingabire W, Hedt-Gauthier B, Nahimana E, Gupta N, Hansen A, Labrecque M, Nkikabahizi F, Mutaganzwa C, Biziyaremye F. A retrospective study of neonatal case management and outcomes in rural Rwanda post implementation of a national neonatal care package for sick and small infants. BMC pediatrics. 2018 Dec;18:1–1.

9. Mwaniki MK, Gatakaa HW, Mturi FN, Chesaro CR, Chuma JM, Peshu NM, Mason L, Kager P, Marsh K, English M, Berkley JA. An increase in the burden of neonatal admissions to a rural district hospital in Kenya over 19 years. BMC Public Health. 2010 Dec;10:1–3.

10. Also U, Gwarzo GD. Morbidity and mortality patterns among neonates in a tertiary hospital. Sahel medical journal. 2020 Jan 1;23(1):47–50.

11. Fajolu IB, Egri-Okwaji MT. Childhood mortality in children’s emergency center of Lagos University Teaching Hospital. Nigerian Journal of Pediatrics. 2011;38(3):131–5.

12. Hoque M, Haaq S, Islam R. Causes of neonatal admissions and deaths at a rural hospital in KwaZulu-Natal, South Africa. Southern African Journal of Epidemiology and Infection. 2011 Jan 1;26(1):26–9.

13. Simpson CD, Ye XY, Hellmann J, Tomlinson C. Trends in cause-specific mortality in an outborn Canadian NICU. Pediatrics. 2010 Dec 1;126(6):e1538–44.

14. Kumar M, Paul VK, Kapoor SK, Anand K, Deorari AK. Neonatal outcomes in a sub-district hospital in North India. Journal of Tropical Pediatrics. 2002 Feb 1;48(1):43–6.

15. Ekwochi U, Ndu IK, Nwokoye IC, Ezenwosu OU, Amadi OF, Osuorah DI. Pattern of morbidity and mortality of newborns admitted to the sick and special care baby unit of Enugu State University Teaching Hospital, Enugu State. Nigerian Journal of Clinical Practice. 2014 May 29;17(3):346–51.

16. Hussain S. Neonatal morbidity and mortality pattern in a tertiary care neonatal unit of a teaching hospital. Am. Ann Pak Inst Med Sci. 2014;10(1):7–11.

17. Islam MN. Neonatal health situation in Bangladesh. The Orion. 2000 May;6:1–4.

18. Rakholia R, Rawat V, Bano M, Singh G. Neonatal morbidity and mortality of sick newborns admitted in a teaching hospital of Uttarakhand. CHRISMED, Journal Health and Research. 2014 Oct 1;1(4):228–34.

19. Shahani Z, Shaikh AR, Gemnani VK, Abro K, Aizuddin AN, Manaf MR, Shahani MP. Neonatal morbidity patterns and admission outcomes: This cross-sectional study was conducted at a tertiary care hospital in Pakistan. Journal of Pharmaceutical Research International. 2022:72–6.

